# Predicting progression to septic shock in the emergency department using an externally generalizable machine learning algorithm

**DOI:** 10.1101/2020.11.02.20224931

**Authors:** Gabriel Wardi, Morgan Carlile, Andre Holder, Supreeth Shashikumar, Stephen R Hayden, Shamim Nemati

## Abstract

**Objective:** Machine-learning (ML) algorithms allow for improved prediction of sepsis syndromes in the ED using data from electronic medical records. Transfer learning, a new subfield of ML, allows for generalizability of an algorithm across clinical sites. We aimed to validate the Artificial Intelligence Sepsis Expert (AISE) for the prediction of delayed septic shock in a cohort of patients treated in the ED and demonstrate the feasibility of transfer learning to improve external validity at a second site.

**Methods:** Observational cohort study utilizing data from over 180,000 patients from two academic medical centers between 2014 and 2019 using multiple definitions of sepsis. The AISE algorithm was trained using 40 input variables at the development site to predict delayed septic shock (occurring greater than 4 hours after ED triage) at varying prediction windows. We then validated the AISE algorithm at a second site using transfer learning to demonstrate generalizability of the algorithm.

**Results:** We identified 9354 patients with severe sepsis of which 723 developed septic shock at least 4 hours after triage. The AISE algorithm demonstrated excellent area under the receiver operating curve (>0.8) at 8 and 12 hours for the prediction of delayed septic shock. Transfer learning significantly improved the test characteristics of the AISE algorithm and yielded comparable performance at the validation site.

**Conclusions:** The AISE algorithm accurately predicted the development of delayed septic shock. The use of transfer learning allowed for significantly improved external validity and generalizability at a second site. Future prospective studies are indicated to evaluate the clinical utility of this model.

## INTRODUCTION

Background: Sepsis remains a significant public health burden with more than 1,700,000 cases diagnosed in the United States each year (1). Mortality associated with sepsis remains high, particularly for those who develop organ failure and shock, despite considerable investment in improving care for these patients. The majority of cases of severe sepsis or septic shock are identified at the time of, or, near the triage time in the Emergency Department (ED). However, approximately 10% of patients with a sepsis syndrome progress to septic shock after triage in the ED (2-4). Prior research has shown that progression to septic shock is associated with worse outcomes (5-7). While significant efforts have been made to determine mortality risk in patients with sepsis in the ED, there has been little investigation into identifying which septic patients will progress to shock (8, 9).

Importance: Patients with delayed onset of septic shock from the ED have up to a 20% higher mortality rate compared to septic patients who do not develop shock or are initially admitted to the ICU (10, 11). Earlier identification of patients at risk for progression to septic shock may thus help appropriate triage and utilization of resources in these patients. We currently lack reliable models to identify patients at high-risk of a delayed progression to septic shock. Prior investigations into this have been limited by small numbers, single-center design, or insufficient test characteristics (3, 12). Machine-learning (ML) techniques allow for a more data-driven and comprehensive approach to diagnosis and prognostication compared to traditional statistical methods. ML methods have been previously used in the detection of sepsis in a variety of clinical scenarios (13-15), with some providing explanations for model outputs; thus ensuring interpretability [15]. Transfer learning, an ML method that allows for fine-tuning (the process of optimizing parameters in a neural network) of a previously trained model during external validation at new site, has the ability to improve site-specific test characteristics from a general model (16) and has not yet been described in the ED.

Goals of This Investigation: The primary objective of this multicenter cohort study was to (1) describe the use of ML techniques to predict the development of delayed septic shock from a cohort of ED septic patients with end-organ damage (defined as severe sepsis according to Center for Medicare and Medicaid Services (CMS), or the “Sepsis 3” international definition). and (2) demonstrate the feasibility of transfer learning to show improved performance and generalizability at a second clinical site.

## METHODS

### Study Design and Setting

This was a retrospective multicenter cohort study of all adult patients (≥18 years old) who were admitted from the emergency department (ED) with sepsis between 2014 and 2019 from 2 large urban academic health centers, the University of California, San Diego, and the Emory University in Atlanta. Institutional review board approval of the study was obtained at both sites with a waiver of informed consent (Emory #110675 and UC San Diego #191098). Emory University has an estimated 192,500 number of annual ED visits, whereas UC San Diego has an estimated 70,000 annual ED visits. Throughout the manuscript we refer to the respective hospital systems as the *development* and the *validation* cohorts. Within in cohort, we refer to two subgroups: a training subgroup and a testing subgroup.

### Definitions and Population

All adult patients who were admitted to the hospital from the two EDs during the study period were evaluated for suspected sepsis via automated query of the electronic medical record (EPIC, Verona, WI and Cerner, Kansas City, MO). Data were abstracted into a clinical data repository (MicroStrategy, Tyson Corner, VA) and included vital signs, laboratory values, sequential-organ failure assessment (SOFA) scores, co-morbidity data (including Charlson comorbidity index scores (CCI)), length of stay, and outcomes. We utilized two definitions of sepsis: the Center for Medicare and Medicine Services criteria for severe sepsis (“CMS severe sepsis”) and the most recent international consensus criteria for assessment of sepsis (“Sepsis-3”) from electronic health records (17-19). We chose to focus on CMS severe sepsis as the primary criteria as this can be calculated in real-time and is a national quality metric. Sepsis-3 data was used for sensitivity analysis. Additionally, we chose “severe sepsis” rather than simple sepsis from the CMS guidelines as patients meeting the criteria have a similar end-organ damage profile to the sepsis definition in Sepsis-3. We defined CMS severe sepsis as the time at which the patient had a culture taken with subsequent antibiotic administration (excluding prophylactic use), the presence of 2/4 systemic inflammatory syndrome (SIRS) criteria, and had evidence of organ dysfunction (e.g., lactate > 2 mmol/L) as defined in the most recent Center for CMS SEP1 guidelines. The onset of CMS severe sepsis (t-sepsis) was the latest timestamp associated with these three events, all of which had to occur within a 6 hours window while the patient was in the ED. Sepsis-3 was defined based on the clinical operationalization of the 2016 international consensus definition as the first of two timepoints: suspected infection and the presence of end-organ damage (18). The suspicion of sepsis (t-suspicion)– was defined as blood cultures and antibiotic initiation (for at least three days, excluding prophylactic use) within 24 or 72 hours, depending on if cultures or antibiotics occur first, respectively. End-organ damage was defined by the 2016 international definition of sepsis as a 2-point increase in the subject’s SOFA score. This included the time of a 2-point increase in the SOFA score (t-SOFA) from up to 24 hours before to up to 12 hours after the t-suspicion (t-SOFA + 24 hr > t-suspicion > t-SOFA – 12 hr). The time of septic shock was defined as the earliest point a patient met criteria for sepsis *and* had use of a titratable vasoactive medication (e.g. norepinephrine).

As with prior research into the identification of patients that develop delayed septic shock following the diagnosis of sepsis, we included patients who developed septic shock between 4 hours after ED triage and up to 48 hours after diagnosis of severe sepsis (8). However, for the purpose of prediction of shock the ML algorithm only made predictions from the time of severe sepsis to time of shock or end of ED visit. We excluded patients that were discharged from the emergency department, patients with an ED length of stay less than 3 hours, and patients who developed septic shock within 4 hours of ED triage. Patients who developed severe sepsis after transferring out of the ED were also excluded.

### Feature Extraction and Machine Learning

Similar to PhysioNet Sepsis Challenge 2019, we included a total of 40 most commonly measured input variables from the electronic health record (EHR) for model development (see Appendix 1) (20). Data sampling began at the time of first measurement in ED. Data was sampled at hourly intervals and handling of multiple measurements within an hour or missing values was performed similar to our previously published work (15). All data used was available to emergency physicians at or ahead of model entry. Values for static demographic information were kept constant for all time bins in an encounter. The median value was used for time-variant predictor features such as vital signs and laboratory tests if a bin contained multiple values. Feature values from prior bins were retained (sample-and-hold interpolation) if data was missing in new bins. Mean values were imputed for features that were missing when there were no prior values. All population-level statistics (e.g., mean values for features that were missing) were calculated on development site training data, and were fixed, then applied to the testing data and validation site data. Appendix 1 includes the frequencies of missing data of input features. For each input feature, we provide the percentage of missingness over a 24 hour time interval, divided into 24 bins, each 1 hour in duration. An input feature (e.g. total bilirubin) checked only once within the first 24 hours would be present 4.1% (1/24) of the time, or alternatively, be missing 95.9% of the time. Prediction of septic shock started at the time CMS severe sepsis was identified. In the analysis using the Sepsis-3 definition of sepsis, prediction of septic shock began at the time patient met criteria for sepsis.

Data was then used to train a modified Weibull-Cox proportional hazards model (AISE, the Artificial Intelligence Sepsis Expert) to predict the onset of septic shock on hourly basis, starting from the time of sepsis up to time of shock. The modification to the AISE model was the inclusion of a two-layer feedforward neural network (of size 10 each) prior to calculations of risk score by the Weibull-Cox model. The neural network was introduced to capture multiplicative risk factors (e.g. age and temperature, where in elderly patients, hypothermia may be more of a risk factor than in young adults). Both the neural network and the AISE parameters were initialized randomly and jointly optimized using an algorithm known as gradient descent (21).

During external evaluation, the pre-trained AISE model derived from data only at the development cohort was fine-tuned on the training subgroup at the validation site, using 20 iterations of the gradient descent algorithm. The AISE model was then evaluated on the testing subgroup of the validation cohort. The purpose of the external evaluation step was to show that the model can be tailored to the characteristics of each local population and provide accurate predictions. This approach falls under the framework of *transfer learning* in machine learning literature and has been shown to improve prediction performance as opposed to a model trained from scratch on external cohorts, in particular when only limited data is available for training (22). In this framework, knowledge gained by solving a classification problem on the development cohort is stored in the weights of the neural network and is carried forward to the target cohort (Figure 1). As such, learning of an accurate and generalizable classifier on the target cohort can be achieved using less training data. This approach has been previously used to fine-tune a neural network model for detection of diabetic retinopathy in retinal fundus photographs (23).

**Figure 1:**
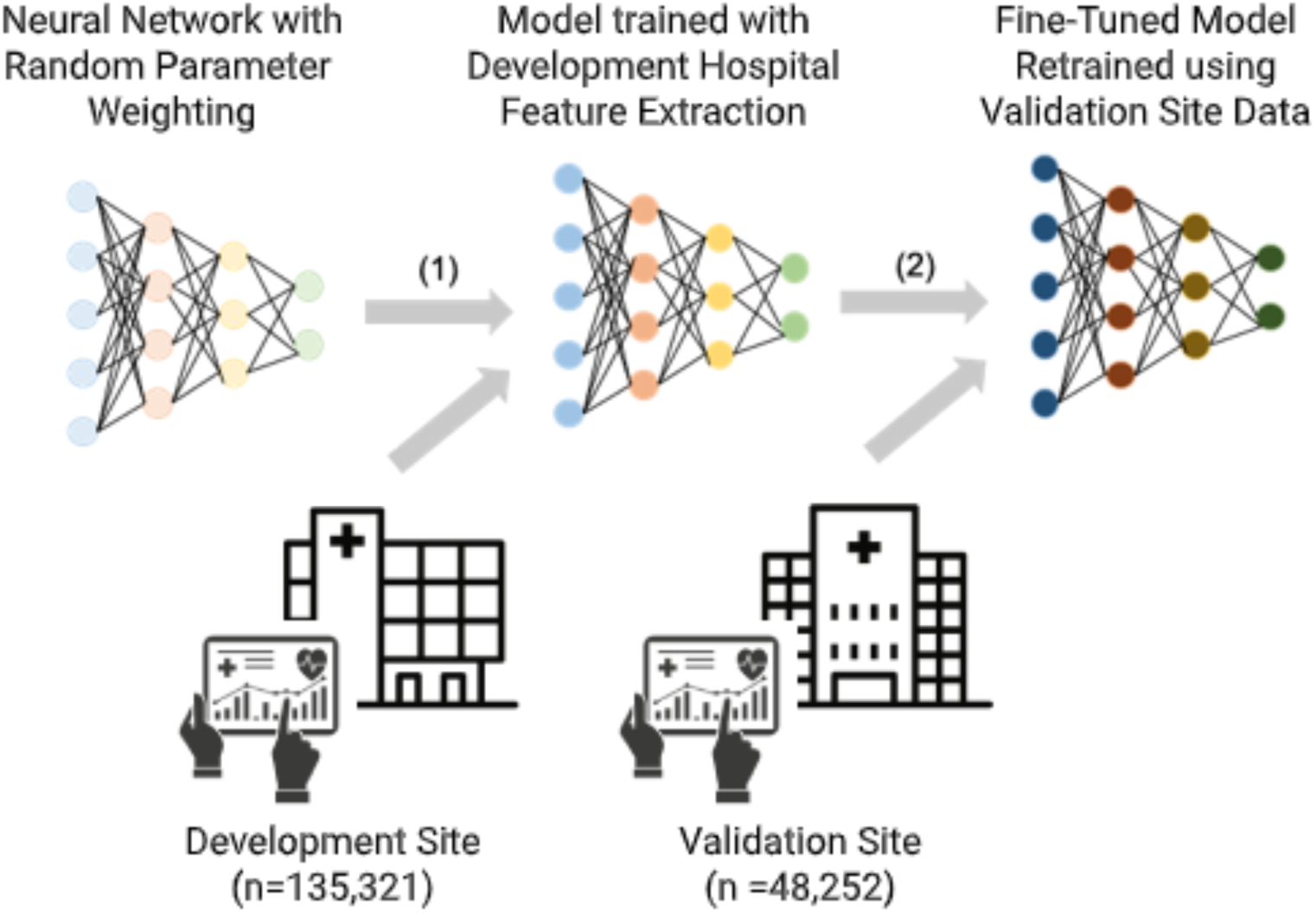
Schematic diagram of transfer learning as implemented in this study. (1) A prediction model neural network (such as AISE) is initialized with random weights and is trained using data from the development site. (2) Starting with a pre-trained model from the development site, the model weights are fine-tuned (or retrained) using relatively small amount of data from the validation site prior to application to the validation cohort. This procedure is called transfer learning.

### Data processing and statistical methods

Features in the development cohort set first underwent normality transformations then were standardized by subtracting the mean and dividing by standard deviation. All the remaining datasets were normalized using the mean and standard deviation computed from the development cohort training set. All continuous variables are reported as medians with 25% and 75% interquartile ranges (IQRs). Binary variables are reported as percentages. We used 10-fold bootstrap cross-validation with an 80%-20% random split within each fold for training and testing subgroups. We report median and interquartile values of the performance statistics, including AUC and specificity (reported at 85% sensitivity) on both the training and testing cohorts. We elected to use a sensitivity of 85% for our model to reflect the need of a screening tool. For the derivation and validation sites, AISE classification for 8, 12, 16, 24 and 36-hour prediction horizons are reported using area under receiver operating characteristic curves (AUCroc) statistics for both the training and the testing folds, as well as specificity (1-false alarm rate).

## RESULTS

### Baseline Characteristics

A total of 135,321 patients were admitted to the hospital from the first hospital system (the *development cohort*) and 48,252 patients were admitted from the second hospital system (the *validation cohort*) site during the study period. Of these, 8,499 patients met criteria for CMS severe sepsis in the development cohort and 6,409 patients in the validation cohort. We identified 643 patients with septic shock in the development cohort and 715 patients with septic shock in the validation cohort, of which 365 (4.3%) and 358 (5.9%) developed septic shock 4 hours after triage and 48 hours after development of severe sepsis, respectively (see Figure 2). Patients in the development cohort who developed delayed septic shock had similar age (66 years old vs 66 years old), had higher SOFA scores (3.8 vs 2.6), and CCI scores (4.0 vs 3.0) than patients that did not develop delayed septic shock (see Table 1). Similar trends were also noted within the validation cohort. The median time from sepsis to the development of shock in the development cohort was 9.9 hours (IQR 5.5 to 18.8 hours) and 7.6 hours (IQR 4.8 to 13.7 hours) in the validation cohort. Inpatient mortality of patients with CMS severe sepsis, but without shock, and those with delayed septic shock in the development cohort were 5.5% and 24.9%, respectively. Likewise, inpatient mortality in the validation cohort with only CMS severe sepsis and those with delayed septic shock were 5.5% and 24.3%. Patients in the development cohort had higher rates of transfers to inpatient hospice care (6.3% and 10.3%) for patients with severe sepsis and delayed septic shock than the patients in the validation cohort (0.7% and 1.7%), respectively. Supplemental Table 1 provides data for similar demographic information using Sepsis 3 definitions at the development and validation cohorts for model development using this definition.

**Table 1:** Demographic comparisons of the derivation and validation sites using CMS severe sepsis definition^a^.

|  | Development site after 4 hours from ED arrival until 48 hours after T0 of severe sepsis |  | Validation site after 4 hours from ED arrival until 48 hours after T0 of severe sepsis |  |
| --- | --- | --- | --- | --- |
| Demographics | Severe sepsis without shock<sup>b</sup> | Severe sepsis with shock<sup>c</sup> | Severe sepsis<sup>b</sup>, Without shock | Severe sepsis with shock<sup>c</sup> |
| Patients, *n* | 4951 | 365 | 4403 | 358 |
| Age, yrs (S.D) | 62 (48, 73) | 65 (54, 74) | 60.5 (47.4, 72.1) | 62.2 (53.1, 72.9) |
| Male, % | 53.2 | 49.3 | 57.2 | 57.3 |
| Race, % |  |  |  |  |
| White | 42.5 | 44.7 | 53.7 | 56.4 |
| Black | 50.9 | 50.7 | 9.5 | 9.8 |
| Asian | 3.2 | 1.6 | 8.9 | 6.9 |
| Other | 3.4 | 3.0 | 27.9 | 26.9 |
| ICU LOS, hrs (IQR) | 0 (0, 46.5) | 95.1 (51.3, 195.5) | 0 (0, 16.5) | 87.9 (46.1, 169.6) |
| ED LOS, hrs (IQR) | 6.5 (5.0, 8.8) | 5.9 (4.6, 7.9) | 9.4 (6.7, 16.1) | 8.2 (6.2, 11.2) |
| CCI, # (IQR) | 4 (2, 6) | 4 (2, 7) | 5 (3, 8) | 5 (3, 8) |
| SOFA, *n* (IQR) | 2.9 (1.8, 4) | 3.8 (2, 5.2) | 1.6 (0.5, 2.8) | 2.8 (3.4, 2) |
| Inpatient mortality, % | 5.5 | 24.9 | 5.5 | 24.3 |
| Transfers to hospice, % | 6.3 | 10.9 | 0.7 | 1.7 |
| Time from ED admission to shock<sup>d</sup>, hrs (IQR) | -- | 11.9 (7.0, 21.0) | -- | 10.0 (6.8, 16.9) |
| Time from sepsis to shock, hrs (IQR) | -- | 9.9 (5.5, 18.8) | -- | 7.6 (4.8, 13.7) |

S.D=standard deviation; yrs=years; LOS=length of stay; ED=Emergency department; IQR=interquartile range; CCI=Charlson comorbidity index; SOFA=sequential organ failure assessment
a- Patients from the overall cohort were included in the study cohorts (derivation and validation cohorts) if they met all of the following criteria: (1) met sepsis criteria according to CMS guidelines<sup>1</sup>, defined as the time at which the patient had a culture taken with antibiotic administration of at least 3 days, AND met SIRS criteria, AND a lactate >2 OR other signs of organ dysfunction, as defined in the CMS guidelines<sup>[18]</sup>; (2) were admitted to the ED for at least 3 hours, and no more than 7 days; (3) developed shock after the first 4 hours of ED admission, if applicable; (4) developed shock after 2 hours of developing sepsis. If applicable.
b- Sepsis is defined as the time at which the patient had a culture taken with antibiotic administration of at least 3 days, AND met SIRS criteria, AND a lactate >2 OR other signs of organ dysfunction, as defined in the CMS guidelines. The time of sepsis onset is the earliest timestamp associated with these events.
c- Septic shock is defined as patients with sepsis who require vasopressor administration.
d- Time until septic shock, defined as the time of vasopressor administration in someone with sepsis

**Figure 2:**
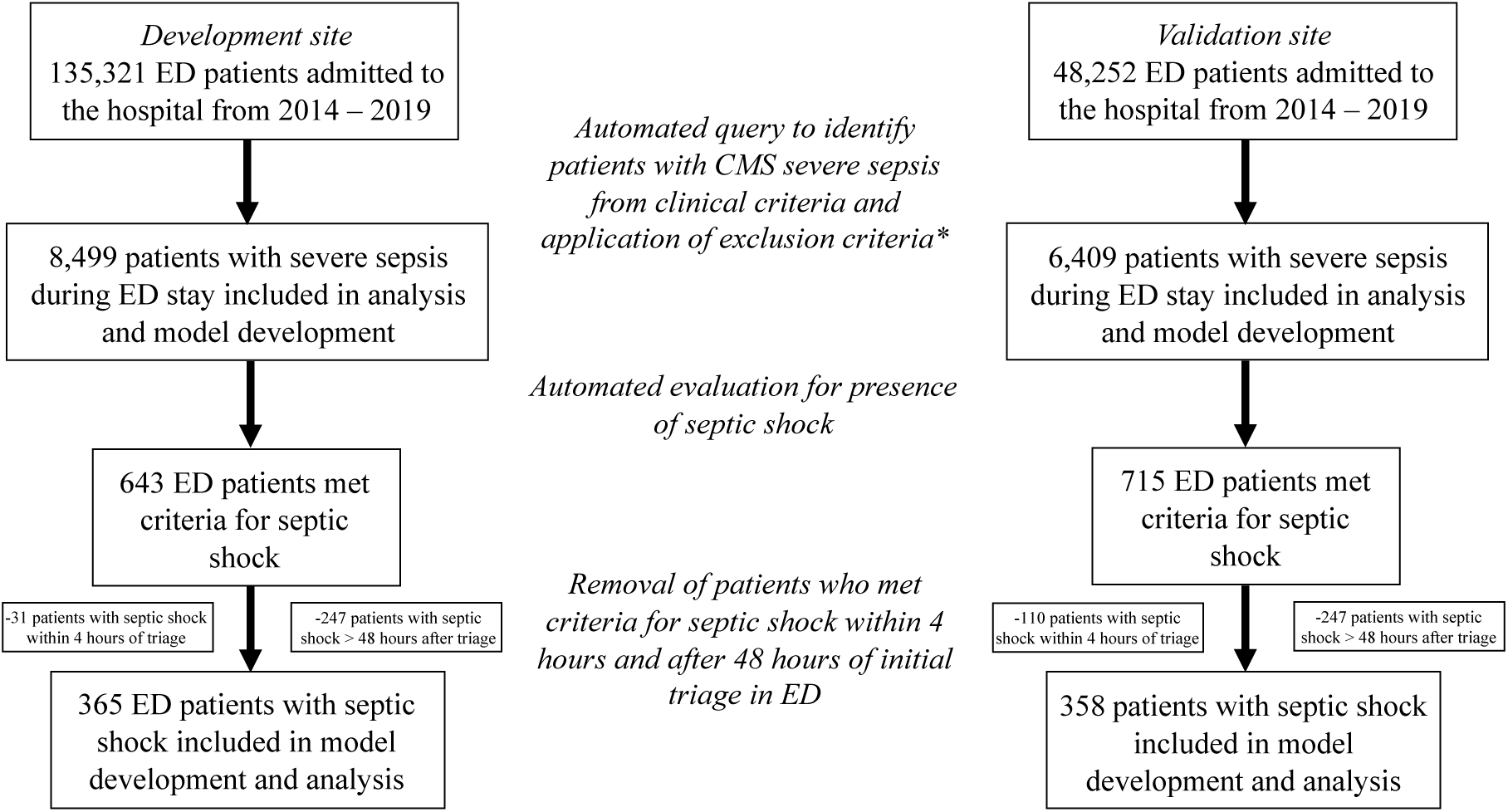
Flowchart describing inclusion of subjects used in the development and validation cohorts using CMS sepsis definitions. * Exclusion criteria: patients that were discharged from the emergency department, patients with an ED length of stay less than 3 hours, patients who developed severe sepsis after transferring out of the ED.

### Performance of AISE for Predicting Delayed Septic Shock

The AUCroc at varying prediction windows after the time of ED sepsis triage to detect delayed septic shock at the development site and validation site is provided in Figure 3. Regardless of definition of sepsis, the AUCroc was 0.80 or higher up to 12 hours in advance in the development cohort. The AUCroc of the ability of the algorithm to predict delayed septic shock at the validation site was > 0.8 at 8, 12 hours using the Sepsis 3 definition and > 0.8 at 8 and 12 hours using the CMS definition. The specificity of the algorithm at varying prediction windows is also provided in Figure 3 for both the development and validation cohorts at 8, 12, 16, 24 and 36 hours. We found that as the prediction window was extended past 16 hours, the AUCroc in both the development and validation cohorts decreased, regardless of definition of sepsis used. Additionally, the AUCroc of the development cohort was greater than that of the validation cohort at almost all prediction windows.

**Figure 3:**
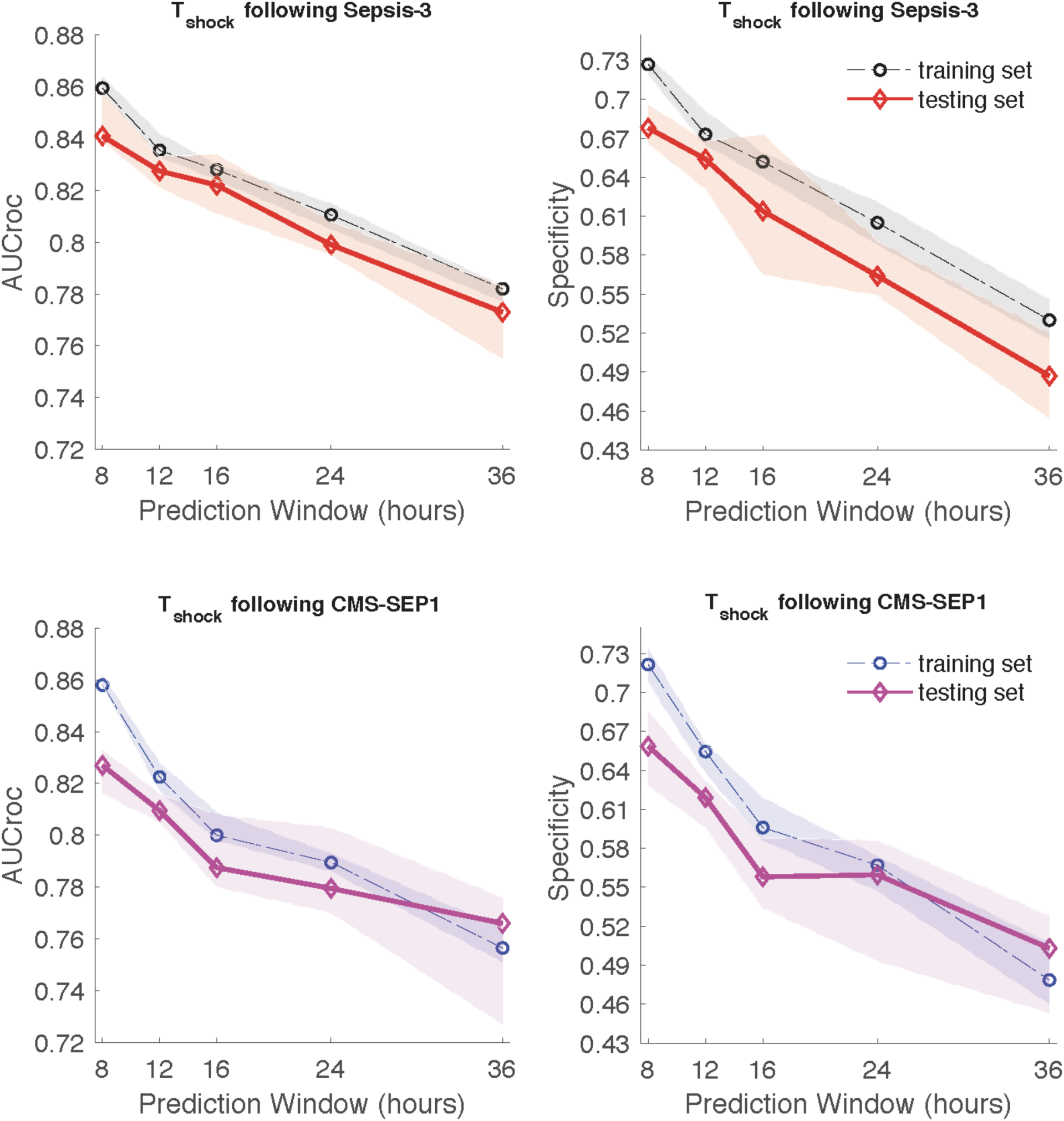
Comparison of AUC and specificity (calculated at 85% sensitivity) of models predicting septic shock at different prediction windows at the validation site. 10-fold cross-validation was performed and a median and inter-quartiles are presented using lines and shaded areas, respectively.

The most weighted input features of the algorithm, in order of importance for the detection of septic shock were systolic blood pressure (SBP), blood urea nitrogen (BUN) level, respiratory rate (RR), temperature and change in blood pressure in the cohort of patients using CMS sepsis definitions. We used a collection of vital sign measurements at a given time (with carry forwarded or sample-and-hold values for missing values) to make a prediction, and then move forward by one hour and make another prediction. Table 2a provides the remainder of the top 20 most important features for the prediction of septic shock in the development cohort and the corresponding change in the AUCroc if removed from the model. In the cohort of patients identified using Sepsis-3, the five most important variables were, in the following order: systolic blood pressure, respiratory rate, heart rate, blood urea nitrogen (BUN), and diastolic blood pressure. Supplemental Table 2 provides the top 20 most important features for the prediction of septic shock and corresponding change in AUCroc if removed from the model.

**Table 2a:** Top 20 most important features (Development Cohort) in septic shock prediction and corresponding change in AUCroc if removed.

|  |  |  |  |
| --- | --- | --- | --- |
| 1 | SBP (0.062) | 11 | Calcium (0.004) |
| 2 | BUN (0.019) | 12 | DBP (0.003) |
| 3 | Resp Rate (0.012) | 13 | Potassium (0.002) |
| 4 | Temp (0.008) | 14 | O2Sat (0.001) |
| 5 | Delta_SBP (0.007) | 15 | Delta_O2Sat (0.001) |
| 6 | Hct (0.007) | 16 | Delta_Temp (0.001) |
| 7 | wbc (0.007) | 17 | MAP (0.001) |
| 8 | Lactate (0.007) | 18 | Delta_EtCO2 (0.001) |
| 9 | Creatinine (0.007) | 19 | Delta_BaseExcess (0.001) |
| 10 | HR (0.006) | 20 | Delta_HCO3 (0.001) |

O<sub>2</sub>Sat=Oxygen Saturation; DBP=Diastolic Blood Pressure; HCO<sub>3</sub>=Bicarbonate; HR= Heart Rate; Temp=Temperature; EtCO<sub>2</sub>=End-Tidal CO<sub>2</sub>; SBP=Systolic blood pressure; The number in parentheses shows the amount of change in AUC if a feature is treated as missing in the model

### Probability of Progression to Septic Shock and Mortality

Supplemental Table 3 provides the mortality rate and percentage of patients who underwent transition to comfort measures based on the algorithm’s prediction for the development of delayed septic shock with a prediction window of 12 hours or less in the development cohort. As the probability of delayed septic shock increased, so did the chance of mortality or transition to hospice, particularly if the probability was greater than 0.8 where combined hospice and mortality rates was 22.6%.

There were only 18 false negative predictions by the algorithm (i.e. patients with septic shock who were miss-classified as not having this), as shown in Supplemental Figure 3. Among these 18 patients, all had probabilities of developing delayed septic shock less than 0.4 and none of the patients died or were transitioned to hospice. In other words, when the algorithm says the patient is not at risk for septic shock there is a very high likelihood that the person is actually not at risk of septic shock and even if misclassified, the patient has a very low chance of mortality. Patients with a false positive (i.e. where the algorithm predicted septic shock, but the patient did not develop this) prediction had a high combined mortality and hospice rate, particularly with a probability between 0.8 and 1.0 (21.3%). So, even if they may not develop shock within the 48 hours window, these patients are more likely to die and merit provider evaluation.

### Test Characteristics of AISE Algorithm at Validation Site After Transfer Learning

The AUCroc in the development cohort at 12 hours in the training and testing subgroups was 0.822 and 0.833 for patients with CMS severe sepsis, respectively (see Figure 4). At the 12-hour prediction window in the validation cohort, prior to transfer learning, the AUC was 0.778 (sensitivity of 0.85, specificity of 0.549) in the cohort of patients with CMS severe sepsis. The positive predictive value of the algorithm is also provided in Figure 4. Negative predictive values of the algorithm for the prediction of delayed septic shock was > 0.99. At the same prediction window and with the same patient population after applying transfer techniques, we found an AUCroc of 0.85 (sensitivity of 0.85, specificity of 0.678) in the testing subgroup at the validation site (DeLong test, p value < 0.001). The five most important features in the algorithm after transfer learning was performed were SBP, HR, temperature, BUN and hematocrit. The remainder of the top 20 most important input features with corresponding AUC change are provided in Table 2b. We then evaluated the AUCroc in the development cohort at a 12-hour prediction window in the training at testing subgroups using Sepsis-3 definitions. Using a 12-hour prediction window, we found the AUCroc in the training subgroup of the validation cohort, prior to transfer learning was 0.792. After the application of transfer learning in the training subgroup, the AUCroc was 0.838 (*p* value from DeLong test < 0.001). In the cohort of patients identified using Sepsis-3, the five most important variables were, in the following order: systolic blood pressure, respiratory rate, heart rate, blood urea nitrogen (BUN), and diastolic blood pressure. Supplemental Table 2 provides the top 20 most important features for the prediction of septic shock and corresponding change in AUCroc if removed from the model. Figure 5 shows the corresponding differences in the AUCroc for prediction of delayed septic shock at 12 hours using the AISE algorithm trained from scratch at the validation site, compared to the algorithm developed using transfer learning with the initial model trained at the development site using an increased numbers of patient encounters in model development.

**Table 2b:**
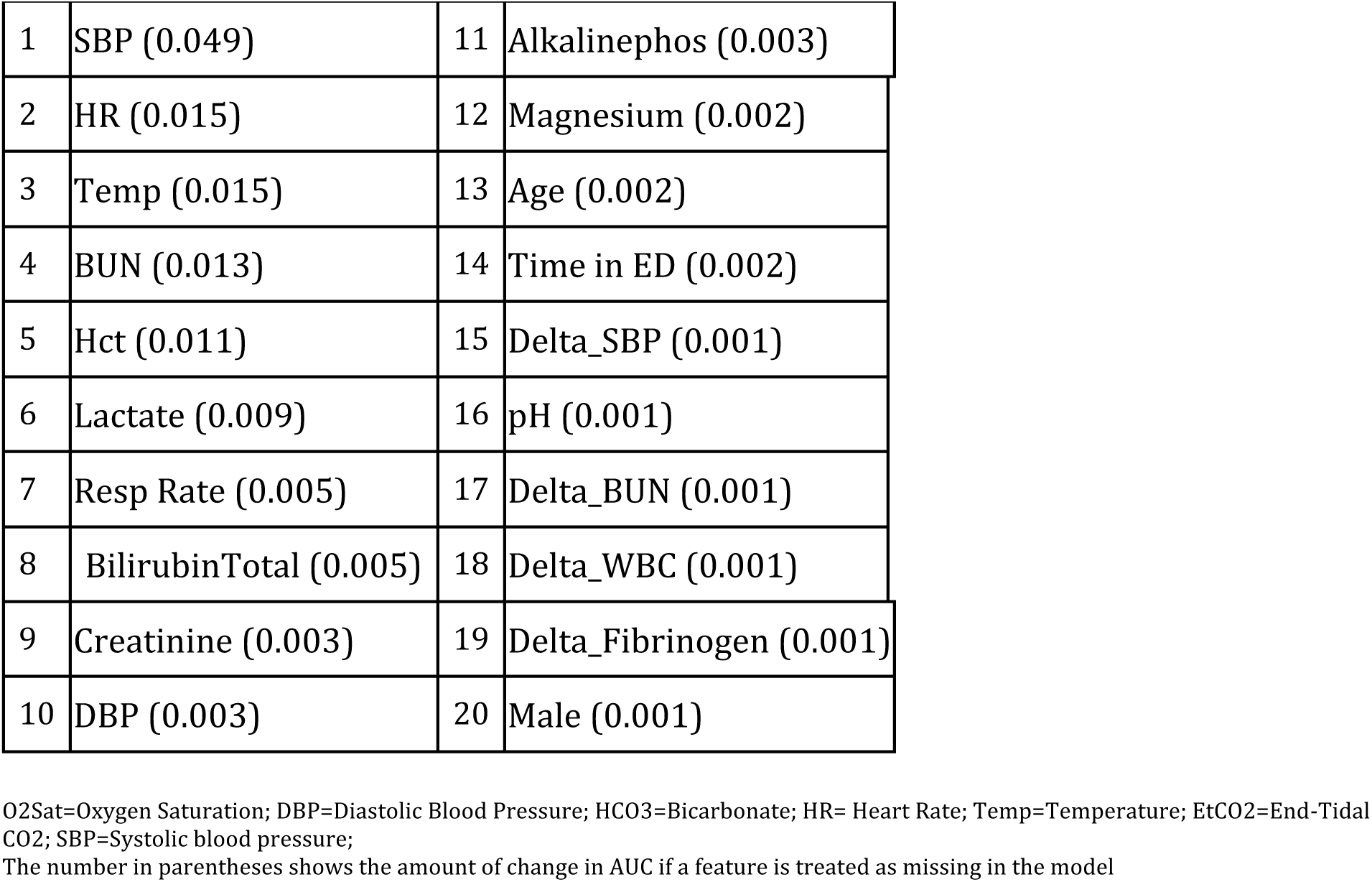
Top 20 most important features (Validation Cohort) in septic shock prediction.

**Figure 4:**
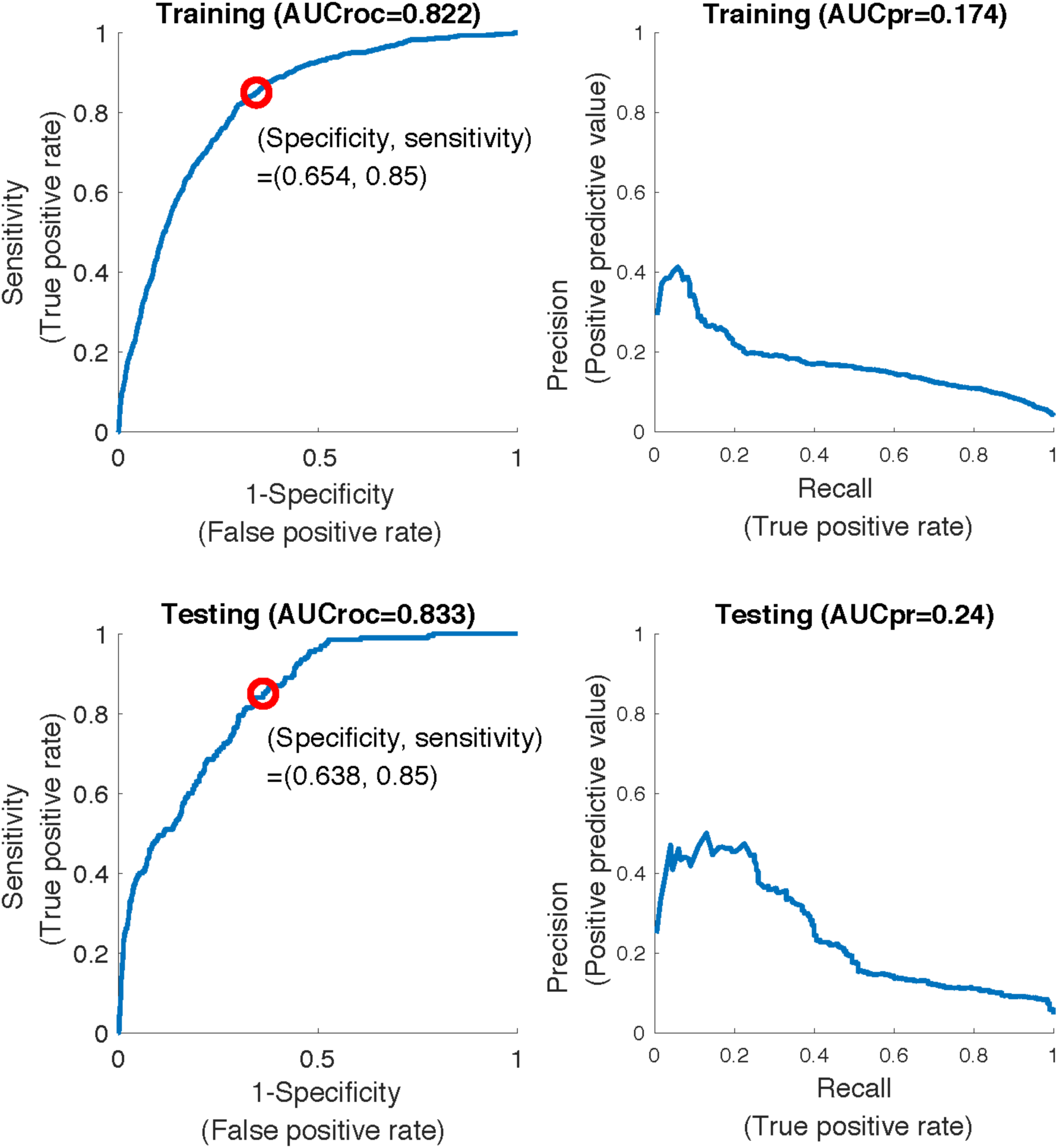
Area under the receiver operating characteristic (AUCROC) and precision-recall curves (AUCPR) for predicting t-sepsis 12 hours in advance at the development site using CMS severe sepsis definition.

**Figure 5:**
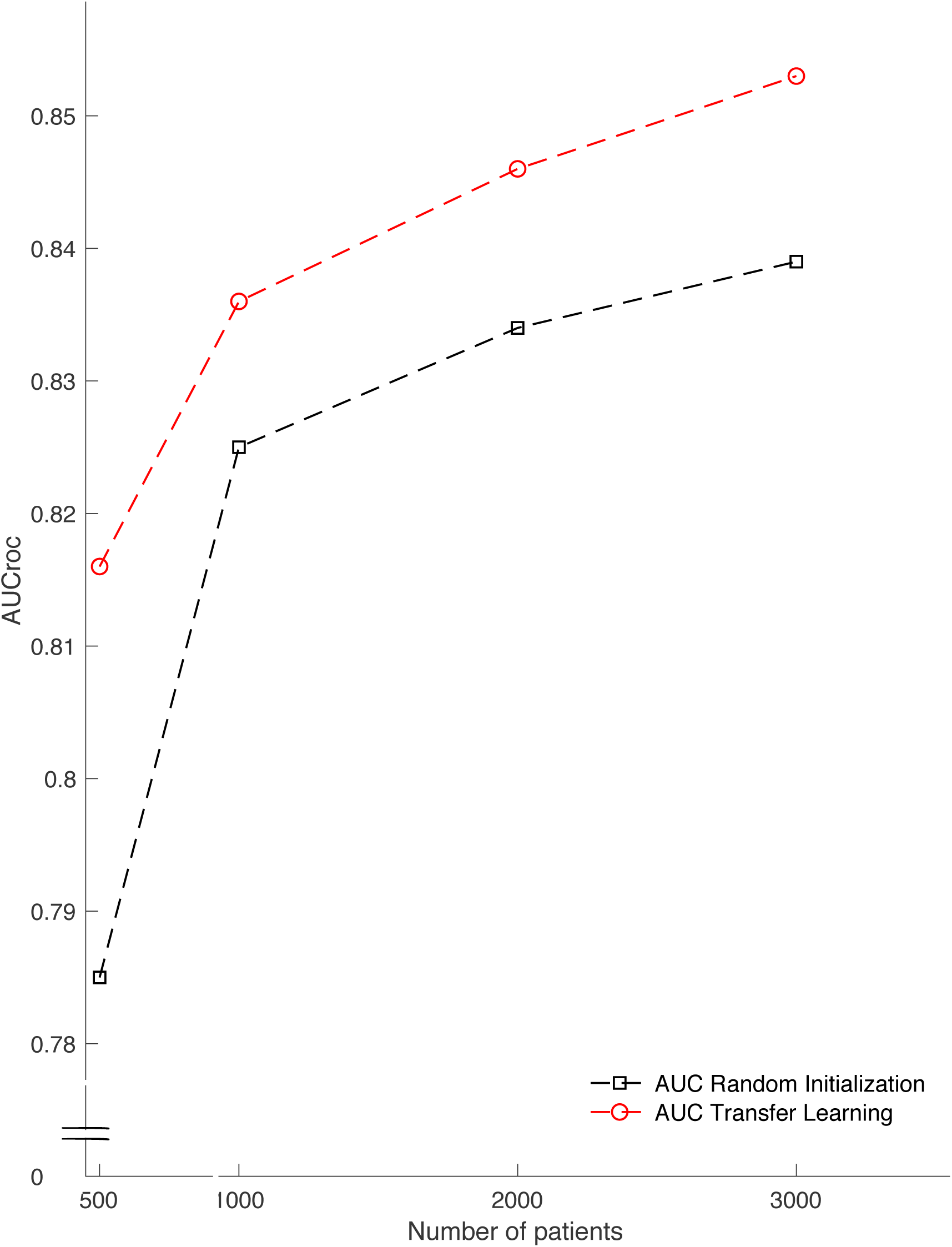
Area under the receiver operating characteristic (AUCROC) of the ability of the AISE algorithm to detect septic shock 12 hours ahead of time in the validation cohort with and without transfer learning (red and black dashed lines, respectively) based on increasing amounts of patient encounters in model development.

## DISCUSSION

We report a retrospective multicenter study that demonstrates the feasibility of a machine-learning algorithm to predict the development of delayed septic shock from 135,321 patients and multiple definitions of sepsis. We then showed that the ML-algorithm can provide excellent predictive characteristics at a separate site via the use of transfer learning. The clinical implications of this are multiple. First, we highlight the need to identify patients at high risk for delayed decompensation as these patients have a significant increase in mortality compared to patients who do not progress to septic shock. We then show favorable test characteristics of ML-algorithms for the identification of patients at high risk of delayed septic shock. Finally, we provide evidence that transfer learning can be used in the ED to boost test characteristics of ML-algorithms at external sites and improve generalizability and portability. These results can help inform the development and implementation of future ML-algorithms to predict the development of conditions or outcomes that are inherently complex and challenging for providers in the ED.

### Detection of Patients at High Risk of Delayed Septic Shock and Associated Mortality

We have shown a significantly higher inpatient mortality in patients with delayed septic shock (24.6%) than patients with CMS severe sepsis but without shock (5.5%). Prior investigations demonstrated similar findings across a variety of conditions (10, 24, 25). While this mortality difference is well described, the ability to identify these patients in the ED has been limited by single-center design and small sample sizes of previous studies. Capp et al. evaluated a cohort of patients (1,336) who progressed to septic shock (111 patients) at least 4 hours after ED arrival and found that female gender, non-persistent hypotension, bandemia, lactate > 4.0 mmol/L and history of coronary artery disease were associated with delayed presentation of septic shock (3). Holder et al. reported that in a cohort of 582 patients, serum albumin < 3.5 g/dL and triage diastolic blood pressure < 52 mm Hg were also associated with decompensation in 108 patients (26). While these findings are important, the clinical utility is challenging to implement as a large majority of patients will meet at least one of the above criteria and will likely not progress to septic shock. Use of early warning systems (e.g. MEWS, NEWS, and VIEWS) for the prediction of adverse events from patients with sepsis have previously been described in the literature((27-30). These scoring systems have performed better than use of quick sequential organ failure assessment (qSOFA) and systemic inflammatory response syndrome (SIRS) for the prediction of adverse events, but their use has been limited by moderate test performance characteristics in septic patients (12).

### Machine-Learning for the Identification of Patients with Delayed Septic Shock

The field of machine learning refers to a subset of artificial intelligence that automates analytical model building to identify patterns in data to predict outcomes. In particular, ML-algorithms are powerful tools for the detection of complicated and non-linear outcomes when traditional statistical methods (e.g. linear regression, recursive partitioning) are overrun by a large number of variables. Recently, ML-algorithms have shown superior test characteristics in the prediction of sepsis in the ED and may lower mortality (13, 14). The AISE algorithm used in this study has previously been shown to have good performance in the detection of sepsis in the ICU at varying prediction windows (4, 6, 8, 12 hours) using 65 readily available variables that yield easily-interpretable scores and can be calculated in real-time (15). Using the same algorithm, but with decreased number of variables for better portability across hospital systems, we found that regardless of the definition of sepsis used, an AUCroc >0.8 for the prediction of delayed septic shock was found for both the development and validation site up to 12 hours in advance. We found the majority of patients who experience a delay in the development of septic shock did so at a median of 7.6 hours (validation site) and 9.9 hours (development site), the majority of which occurred after transferring out of the ED.

Given the excellent test characteristics of the AISE algorithm at these time points, the utility of our findings is clinically relevant, as implementation may decrease unanticipated ICU transfers, prompt a change in management (e.g. fluids or broadening of antibiotics), and raise situational awareness of providers for these high-risk patients. Importantly, we also found that the false positive predictions by the AISE algorithm (those predicted to develop septic shock, but did not) had mortality and transition to hospice rates similar to true positives. This was maintained across sites that had a significant difference in the proportion of septic patients transferred to hospice care. While inappropriate identification for delayed septic shock worsens test characteristics of the AISE algorithm, these patients likely benefit from additional evaluation and potential interventions to prevent decompensation and potentially death.

### Feasibility and Performance of Transfer Learning

Transfer learning is a technique used in machine learning to translate patterns and extracted features learned in one setting and generalizing those patterns in another setting. This allows for ML models developed from a large robust data set to be fine-tuned onto a more sparse data set (31). We found that the use of transfer learning techniques dramatically improved the AUCroc for the prediction of delayed septic shock at a hospital system geographically distinct and consisting of different patient populations with an algorithm trained at an external site (see Figure 1). While others have demonstrated similar results in medical applications of ML, to our knowledge, this is the first instance of a ML-algorithm using transfer learning for an application in the ED (32-34). Previously, decision rules used in emergency medicine have shown impressive test characteristics at the institutions or regions that developed them but attempts at external validation have yielded lower sensitivity and specificity, limiting general use (35, 36). Transfer learning techniques are positioned to further adoption and generalizability of prediction rules and ML-algorithms in external environments, allowing portability of an algorithm from site to site while maintaining superior test characteristics that can be available in real-time to providers (31, 37, 38)

Transfer learning is best applied when a source data set features and outcomes of interest are generalizable, as are the variables used by the AISE algorithm. Whereas some emphasis in machine learning has been focused on knowledge distillation from large implemented ML-algorithms to simpler models that can be applied broadly (for example, a rule-based decision tree), transfer learning techniques allow complicated models such as the one evaluated in this study to be applied to clinical scenarios in more resource limited environments with smaller target data sizes. One strong advantage of this particular implementation and algorithm is privacy and data security. Whereas previously reported methods co-mingle data from multiple institutions to yield stronger test characteristics, this approach may infringe upon a covered entity’s autonomy over protected healthcare data and terms of patient data privacy (39-41). We report a ML process that uses transfer learning in such a way that affords hospital systems the ability to maintain data privacy and does not require a data transfer to a central location for fine-tuning, but rather can be completed at an individual site.

### Limitations

We report results from a single validation site and while we feel confident in the use of transfer-learning to improve external validity, results should be tested on a more geographically diverse patient population. The definitions for sepsis, severe sepsis and septic shock used in this manuscript are based on previously described criteria from CMS and international guidelines and have been optimized for analysis of large clinical databases, but accurate diagnosis of sepsis often requires clinical chart-view and adjudication by multiple reviewers (42). Additionally, we did not include patients who had simple sepsis (e.g. 2/4 SIRS + suspected infection) without any end-organ damage and thus may have not included some patients who developed delayed septic shock without preceding severe sepsis. Patient populations and the diagnostic approach to sepsis may change in the coming years and such algorithms may require revisions, although the proposed transfer-learning framework is well-suited for model fine-tuning, and this form the basis of future studies. Further, we limited the number of input features to 40 most commonly measured vitals and labs. While this was done to increase portability of the algorithm and allow for seamless EMR-agnostic deployment via Health Level Seven (HL7) standard protocols, we acknowledge there are additional variables that could be used to improve the prediction of delayed septic shock. Smaller clinical sites may lack experience or the infrastructure at this point for implementation of such algorithms. However, major EHR distributors are interested in such technological advances and may make implementation easier for interested sites. As we have shown in Figure 5, clinical sites with fewer patients benefit more from model development with transfer learning. Finally, application of this study to broader clinical use will require further validation with prospective, randomized clinical trials assessing patient-centered outcomes (43).

## Conclusion

Patients who develop septic shock 4 hours after triage in the ED have in-hospital mortality approximately five times than those who do not progress to septic shock. The AISE algorithm was trained to provide excellent AUCroc at identifying patients at risk of delayed development of septic shock, particularly at prediction windows of 8 to 12 hours. The most important features in the detection of delayed septic shock with systolic blood pressure, blood urea nitrogen, respiratory rate, temperature and change in blood pressure. Transfer learning was effective at augmenting prediction performance at a second, distinct clinical site. Future prospective studies are required to validate the use of such techniques in clinical practice.

## Data Availability

Access to the computer code used in this research is available upon request to the corresponding author.

---

**Supplemental Table 1:** Demographic comparisons of the derivation and validation sites using sepsis-3 criteria.

|  | Development site |  | Validation site |  |
| --- | --- | --- | --- | --- |
| Demographics | Sepsis<sup>a</sup>,<br>no shock | Sepsis<br>with shock<sup>c</sup> | Sepsis<sup>a</sup>,<br>no shock | Sepsis<br>with shock<sup>c</sup> |
| Patients, *n* (%) | 10087 | 522 | 4984 | 465 |
| Age, yrs (S.D) | 66 (53, 78) | 66 (55, 75) | 62.9 (51.6, 74.2) | 62.3 (53.5, 73.7) |
| Male, % | 53.1 | 49.4 | 59.2) | 60.6 |
| Race, % |  |  |  |  |
| White | 48.1 | 48.6 | 51.9 | 56.3 |
| Black | 45.9 | 46.1 | 9.2 | 9.2 |
| Asian | 2.9 | 1.4 | 8.8 | 5.2 |
| Other | 3.1 | 3.9 | 30.1 | 29.3 |
| ICU LOS, hrs (IQR) | 0 (0, 38.6) | 95.8 (51.8, 198.6) | 0 (0, 18.3) | 93.9 (51.0, 200.9) |
| CCI, # (IQR) | 3 (2, 5) | 4 (2, 7) | 5 (3, 9) | 5 (3, 9) |
| SOFA, *n* (IQR) | 2.6 (2.0, 4.0) | 3.8 (2, 5.1) | 2.3 (1.6, 3.5) | 3.6 (2.3, 5.3) |
| Inpatient mortality, % | 4.2 | 24.9 | 6.3 | 23.4 |
| Transfers to hospice, % | 5.6 | 12.3 | 0.8 | 1.8 |
| Time from ED admission to shock<sup>d</sup>,hrs (IQR) | -- | 11.2 (6.8, 20.81) | -- | 9.9 (6, 17.9) |
| Time from sepsis to shock, hrs (IQR) | -- | 9.5 (5.6, 19.3) | -- | 8.8 (5.5, 16.7) |

S.D=standard deviation; yrs=years; LOS=length of stay; ED=Emergency department; IQR=interquartile range; CCI=Charlson comorbidity index; SOFA=sequential organ failure assessment
a- Patients from the overall cohort were included in the study cohorts (derivation and validation cohorts) if they met all of the following criteria: (1) met sepsis criteria according to sepsis-3 criteria [17], defined as the first of two timepoints: suspicion of infection or t-suspicion (blood cultures and antibiotics within 24 or 72 hours, depending on if cultures or antibiotics occur first, respectively), and the time of a 2-point increase in the Sequential Organ Failure Assessment (SOFA) score (t-SOFA) from up to 24 hours before to up to 12 hours after the t-suspicion (t SOFA + 24 hr > t suspicion > t-SOFA - 12 hr); (2) were admitted to the ED for at least 3 hours, and no more than 7 days; (3) developed shock after the first 4 hours of ED admission, if applicable; and (4) developed shock after 2 hours of developing sepsis. If applicable.
b- Sepsis is defined as the first of two timepoints: suspicion of infection or t-suspicion (blood cultures and antibiotics within 24 or 72 hours, depending on if cultures or antibiotics occur first, respectively), and the time of a 2-point increase in the Sequential Organ Failure Assessment (SOFA) score (t-SOFA) from up to 24 hours before to up to 12 hours after the t-suspicion (t SOFA + 24 hr > t suspicion > t-SOFA - 12 hr); (2) were admitted to the ICU for at least 8 hours, and no more than 20 days; and (3) developed shock after the first 4 hours of ICU admission, if applicable.
c- Septic shock is defined as patients with sepsis who require vasopressor administration.
d- Time until septic shock, defined as the time of vasopressor administration in someone with sepsis

**Supplemental Table 2.** Top 20 most important features (Development Cohort) in septic shock prediction at 12 hours and corresponding change in AUCroc if removed using Sepsis 3 definition.

|  |  |  |  |
| --- | --- | --- | --- |
| 1 | SBP (0.050) | 11 | MAP (0.002) |
| 2 | Resp Rate (0.30) | 12 | Delta_Potassium (0.002) |
| 3 | HR (0.021) | 13 | Delta_MAP (0.001) |
| 4 | BUN (0.008) | 14 | Delta_Resp (0.001) |
| 5 | DBP (0.004) | 15 | Delta EtCO2 (0.001) |
| 6 | FiO2 (0.003) | 16 | Delta_BaseExcess (0.001) |
| 7 | Magnesium (0.003) | 17 | Delta_HCO3 (0.001) |
| 8 | Delta_Magnesium (0.003) | 18 | Delta_FiO2 (0.001) |
| 9 | Potassium (0.003) | 19 | pH (0.001) |
| 10 | Temperature (0.002) | 20 | AST |

SBP = systolic blood pressure; HR = heart rate; BUN = blood urea nitrogen; DBP diastolic blood pressure; MAP = mean arterial pressure; AST = aspartate aminotransferase

**Supplemental Table 3.**
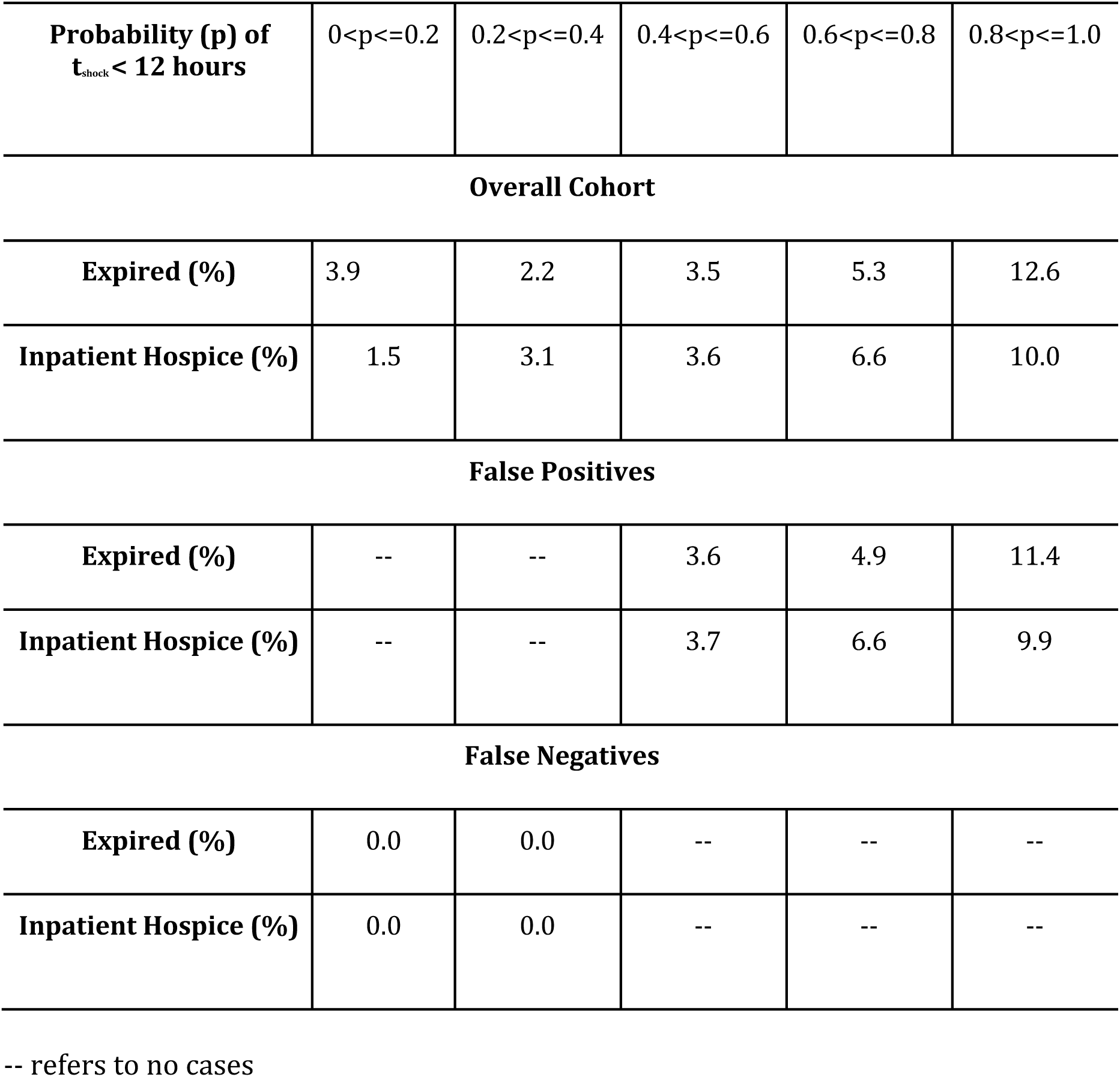
Relationship between probability of septic shock and mortality for the entire cohort, among the false positive cases, and the false negative cases.

| **Probability (p) of**<br>**t<sub>shock</sub> < 12 hours** | 0<p<=0.2 | 0.2<p<=0.4 | 0.4<p<=0.6 | 0.6<p<=0.8 | 0.8<p<=1.0 |
| --- | --- | --- | --- | --- | --- |
| **Overall Cohort** |  |  |  |  |  |
| **Expired (%)** | 3.9 | 2.2 | 3.5 | 5.3 | 12.6 |
| **Inpatient Hospice (%)** | 1.5 | 3.1 | 3.6 | 6.6 | 10.0 |
| **False Positives** |  |  |  |  |  |
| **Expired (%)** | -- | -- | 3.6 | 4.9 | 11.4 |
| **Inpatient Hospice (%)** | -- | -- | 3.7 | 6.6 | 9.9 |
| **False Negatives** |  |  |  |  |  |
| **Expired (%)** | 0.0 | 0.0 | -- | -- | -- |
| **Inpatient Hospice (%)** | 0.0 | 0.0 | -- | -- | -- |

-- refers to no cases

